# Understanding Patient Perceptions of Genetic Testing to Predict Type 2 Diabetes Risk After Gestational Diabetes

**DOI:** 10.1101/2025.11.02.25339227

**Authors:** Ria Patel, Martha Christodoulou, Zoe Taylor, Prajwal Shetty, Julia Zöllner

**Author notes:** **Corresponding author:** Dr Julia Zollner.

## Abstract

**Aims:** Women with gestational diabetes mellitus (GDM) face increased lifetime risk of type 2 diabetes (T2DM). Genetic risk-predictive testing could help identify those at highest risk and guide preventative care. We aimed to assess perceptions of genetic risk scores to help inform future implementation.

**Methods:** An online survey of 112 women with current or prior GDM assessed willingness for genetic and non-genetic risk testing, attitudes lifestyle motivation, and data-use concerns. Quantitative analyses were complemented by thematic analysis of free-text responses.

**Results:** Overall, willingness was high for both genetic testing (83.9%) and non- genetic (90.2%), with no significant difference between them (p = 0.083). Participants identifying as White reported greater willingness for genetic testing (p = 0.020) and stronger agreement that testing should be available on the NHS (p = 0.032) than N=non-White participants. Attitudes toward genetic testing were positive and associated with both willingness to test and support for NHS availability (p < 0.001). Younger participants were more motivated to modify lifestyle behaviours (p = 0.015). Overall, concerns around data collection were low; although free-text responses highlighted health insurance implications, psychological burden, actionability of results, and timing of testing as salient themes.

**Conclusions:** Women with GDM were receptive to genetic risk-prediction for T2DM, with low concerns around data usage. Demographic differences in acceptability and motivation highlight the need for inclusive, targeted communications and lifestyle support alongside integration testing into postnatal-GDM care.

**What is already known?:** We know that women with Gestational Diabetes Mellitus (GDM) have an increased risk of developing Type 2 Diabetes Mellitus (T2DM) later in life. Genetic risk scores can stratify women by their likelihood of developing T2DM following GDM. This risk information could help to inform women’s lifestyle choices and help prevent progression to T2DM. However, evidence is limited regarding women’s willingness to undergo genetic testing, how risk information might shape lifestyle behaviours, and their concerns about data collection and privacy.

**What this study has found?:** Our study found that our participants with current or previous GDM were fairly receptive to genetic testing to predict their future risk of T2DM. There was no significant difference between reported acceptability of a non-genetic test and a genetic test. Participants held positive attitudes towards genetic testing and fairly low concerns about data use and privacy. However, the acceptability of this testing and motivation for lifestyle changes varied by participant demographics (including age, education and ethnicity). Additionally, free-text responses identified concerns over implications for health insurance, the timing of testing, psychological burden of knowing results and the actionability of results.

**What are the implications of the study?:** To support equitable engagement and uptake of genetic risk-prediction testing, inclusive education and communication strategies are needed - particularly on the actionability of results and data collection/storage policies. Although not directly measured, our findings point to the value of culturally responsive, trust-building communication—delivered with cultural humility and empathy—to address uncertainties and enable informed choice. These findings support the use of genetic predictive testing within postnatal GDM care in addition to targeted interventions to support lifestyle changes and prevent progression to type 2 diabetes.

## Introduction

In the UK, gestational diabetes mellitus (GDM) affects approximately 10-20% (1). Women with a history of GDM face a markedly increased risk of developing type 2 diabetes mellitus (T2DM), with estimates suggesting up to 70% will develop T2DM within 10 years postpartum (2, 3). Despite this elevated risk, engagement with postnatal screening and lifestyle interventions to prevent T2DM remains suboptimal with many researchers calling for strategies to increase screening uptake (4, 5). Early identification of women at greatest risk could enable more targeted prevention strategies and improve long-term outcomes (5).

Advances in genomics have made it possible to assess an individual’s genetic risk of progression to T2DM following GDM, complementing traditional clinical risk factors used in risk scores such as body mass index, glucose tolerance, and family history (6). Genetic testing and risk stratification tools could inform targeted interventions, promote lifestyle modification, and optimise follow-up care for these patients (6). However, the successful implementation of this genetic testing in clinical settings depends not only on its scientific validity but also on patient and public understanding and acceptance. Despite increasing research into public and patient attitudes toward genomic testing, relatively little is known about how women with GDM perceive such testing to inform their overall risk of T2DM. However, concerns frequently arise regarding the potential misuse of genetic information, data privacy, and the burden of risk information (7). Moreover, sociodemographic differences, including ethnicity, education, and age, have been shown to influence acceptability and perceived utility of genetic testing (8). Understanding women’s willingness to undergo this genetic testing, their concerns surrounding this, and any factors influencing these attitudes is crucial to integrating genomics into diabetes prevention and postnatal care.

Previous studies examining attitudes toward genetic testing in pregnancy and reproductive health contexts have generally reported positive views, with participants valuing the knowledge gained from testing, particularly when the outcome tested for is perceived as preventable, such as T2DM (9, 10). However, motivation to act on risk information represents another critical consideration. While genetic testing may raise awareness of personal risk, it does not necessarily translate into behaviour change as some individuals may feel overwhelmed when faced with this test result (11). Previous research suggests that risk communication can both empower and overwhelm individuals, depending on their understanding and perceived control over the condition (12). For women with GDM, balancing the demands of motherhood with preventive health behaviours may present additional barriers (13). Exploring how women anticipate responding to risk information—through diet, exercise, or preventative medicines—can therefore help identify opportunities for supportive interventions to accompany genetic testing.

This study aimed to explore the acceptability of, and attitudes toward, genetic testing to assess future risk of T2DM among women with current or previous GDM across the UK. Specifically, it sought to (1) compare willingness to undergo genetic and non-genetic testing; (2) examine associations between attitudes towards genetic testing, demographic factors, and willingness to test; (3) assess participants’ motivation to modify lifestyle behaviours in response to potential risk information; and (4) identify key concerns related to the collection/use of genetic data. By integrating quantitative and qualitative analysis, this study provides an in-depth understanding of women’s perspectives to help inform the implementation of genetic testing in post-GDM care.

## Methods

### Survey design and piloting

The survey aimed to explore women’s perceptions regarding genetic testing to assess future T2DM risk following their experience of GDM across the UK. The survey consisted of 17 closed and open-ended questions (Appendix A). Participants were asked about their willingness to undergo non-genetic and genetic testing, views on the availability of such testing through the NHS, and motivation to modify lifestyle behaviours (diet, exercise, or preventative medication use) based on potential risk information. Attitudes toward genetic testing were measured using a validated scale (14, 15). Additional bespoke items assessed overall perceptions of medical research and concerns regarding the potential use, privacy, and security of genetic data. Demographic information, including age, ethnicity, and education was also collected.

An initial draft of the survey was reviewed through patient and public involvement and engagement (PPIE) by a public contributor with lived experience of gestational diabetes, and by an interdisciplinary, cross-university panel of researchers with expertise in gestational diabetes. We conducted a small pilot with an ethnically diverse group of participants to gather one-to-one feedback on the draft survey. Based on their feedback, we refined items for readability (lay terms), brevity, and usability, and adjusted the order/skip patterns. The pilot formed part of our PPI process.

### Participants and Survey distribution

Eligible participants in this study were women based in the UK with current or previous experience of gestational diabetes mellitus. Data collection occurred from the 1^st^ April 2025 to the 16^th^ September 2025. We distributed the survey via multiple channels, using a snowball sampling approach to extend reach through participant and partner networks. Channels included Gestational Diabetes UK, Diabetes UK, Diabetes Research & Wellness Foundation, the ICP Support charity for intrahepatic cholestasis of pregnancy, local maternity support groups, and online forums such as Mumsnet.

### Data Analysis

Data was analysed using IBM SPSS Statistics software (Version 28). Due to small counts within demographic groups, we collapsed two variables for analysis. Ethnicity was grouped into White and Non-White. Education was grouped into low (no formal qualifications/GCSE’s [equivalent of finishing school at 16 years]), medium (A levels/BTEC [equivalent of finishing school at 18 years]), and high (University degrees).

Descriptive statistics were used to summarise participant characteristics and responses, including means and standard deviations for continuous variables and frequencies and percentages for categorical variables. Differences in paired responses i.e. willingness to take non-genetic and genetic tests were examined using the Related-Samples Wilcoxon Signed Rank Test. Associations between categorical demographic variables (e.g., ethnicity, education level, previous research experience) and ordinal outcome variables were assessed using the Kruskal–Wallis Test for independent samples. Where significant Kruskal–Wallis results were found, post hoc pairwise comparisons were conducted. Associations between continuous or ordinal variables (e.g. attitude scores, concern scores, age) were examined using

Kendall’s tau-b correlation. Statistical significance was set at *p* < 0.05 for all tests. Given the large number of comparisons (N=29), the Bonferroni adjustment was used to control for the risk of Type I error. Analyses were grouped into four families of hypotheses:

1. Willingness (6 tests, αadj=0.0083) and Agreement measures (3 tests, αadj=0.0167)
2. Attitudes towards testing (5 tests, αadj=0.010)
3. Lifestyle influence (3 tests, αadj = 0.0167)
4. Motivation (12 tests, αadj=0.0042).

We report uncorrected p-values and indicate which findings remain significant under the family-wise Bonferroni-adjusted thresholds. For significant ANOVA tests we ran Tukey’s HSD for pairwise comparisons. As these are already adjusted for multiple pairwise comparisons, they were not included in any Bonferroni correction.

Open-ended responses to free text questions related to participants’ concerns and perceptions of genetic testing were analysed thematically using an inductive approach to identify key themes and subthemes (16).

### Ethics approval

This study was approved by the Institute for Women’s Health (IfWH) low risk ethic committee (LREC) (ID: IfWH_LREC_003_2024_25) based at University College London. Completion of the survey was taken as informed consent to participate.

## Results

In total, 129 participants consented to take part in the survey by submitting responses. Of these, 6 reported no experience of gestational diabetes mellitus, and 11 were excluded due to incomplete data (only the first question was answered). This left 112 responses for analysis. Participant demographics are presented in Table 1. Participants most commonly identified as White or White British (46.6%), with responses spanning a range of other ethnicities. Postcode data demonstrated responses from a wide geographical region across the UK.

**Table 1.**
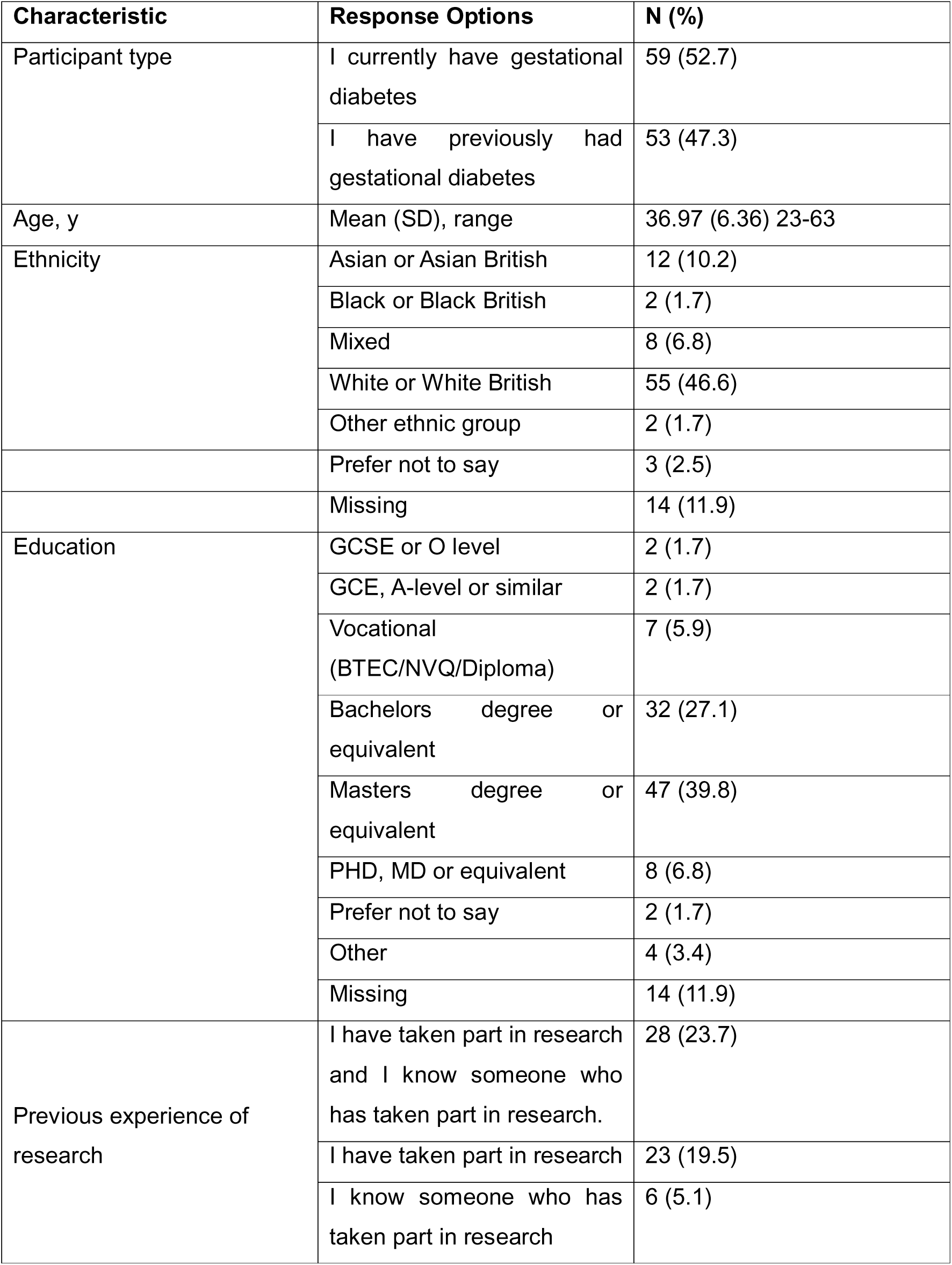
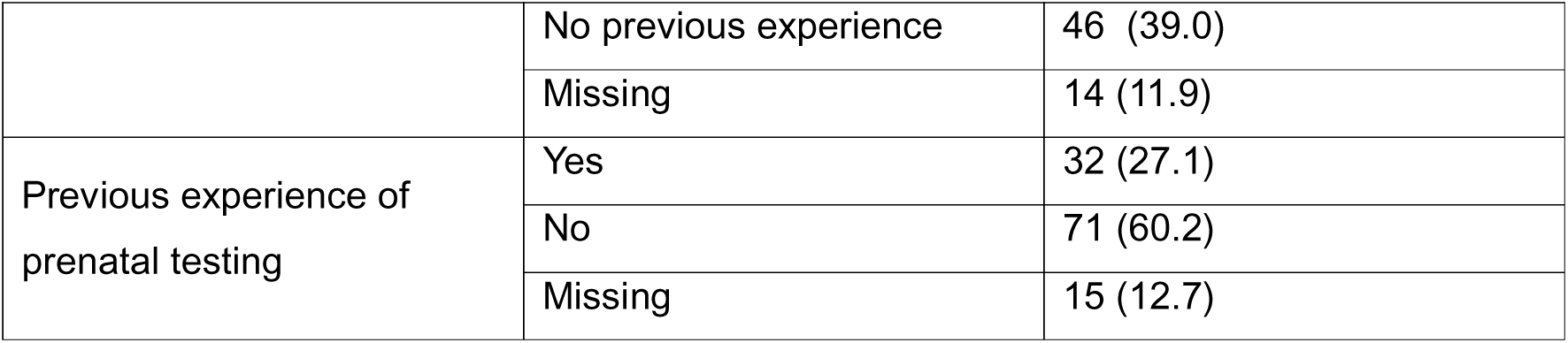
Participant Demographics.

### Acceptability of non-genetic and genetic testing

Participants were asked how willing they would be to take a simple test (not based on genetics) to find out their chance of developing diabetes in the future. 101 participants (90.2%) indicated they were willing to take this test, compared to 1 (0.9%) who was unwilling and 10 (8.9%) who were unsure.

Participants were also asked how willing they would be to take a simple genetic test (a blood test) to find out their chance of developing diabetes in the future. 94 participants (83.9%) indicated they would be willing, compared to 4 (3.6%) who were unwilling and 14 (12.5%) who were unsure. There was no statistically significant difference between participants’ willingness to take a simple non genetics-based test and their willingness to take a genetics-based test, suggesting participants expressed similar levels of willingness to take both tests (*Z* = 1.73, *p* = 0.083).

Difference in willingness to take the non-genetic test was not statistically significant across White and non-White participants (*H*(1) = 3.05, *p* = 0.081), suggesting comparable willingness across these two groups (Mean Rank: White = 50.40, Non-White = 44.50). However, willingness to undergo a genetic test for diabetes risk differed significantly across these two groups (*H*(1) = 5.43, *p* = 0.020), with White participants (Mean Rank = 51.33) showing higher willingness than Non-White participants (Mean Rank = 41.50). There was no significant difference in willingness to take a non-genetics or genetics test across education groups (p=0.210 and p=0.230 respectively), and no significant relationship between willingness and participants’ age (p=0.129).

Participants were asked to what extent they agree or disagree that genetic testing to assess future risk of type 2 diabetes following gestational diabetes should be available on the NHS. 65 (58%) indicated that they strongly agree, 35 (31.3%) indicated they agree, 11 (9.8%) neither agreed nor disagreed and 1 (0.9%) strongly disagreed.

There was no significant association between the extent to which participants agreed that this testing should be available and their education group or age (p=0.764 and p=0.663 respectively). However, there was a statistically significant difference between White and Non-White participants (*H*(1) = 4.62, *p* = 0.032), with White participants (Mean Rank = 51.97) agreeing more strongly that genetics-based testing to assess future diabetes risk should be available on the NHS compared with Non-White participants (Mean Rank = 39.46).

After Bonferroni adjustment, (αadj= 0.0083 for willingness tests and αadj = 0.0167 for agreement tests), none of the reported differences remained statistically significant.

### Attitudes towards genetic testing and research

Attitudes towards genetic testing were positive, with a mean attitude score of 16.85 (SD= 3.93, range 4-20), where higher scores indicating better attitudes towards genetic testing. Attitude scores did not significantly differ across White and Non-White participant groups (p=0.311), education groups (p=0.128) and attitude scores were not significantly associated with age (p=0.899), suggesting that overall attitudes towards genetic testing were positive and consistent across participant demographics. Participants with higher attitude scores were significantly more likely to indicate that they would be willing to have a genetic test to assess their future risk of type 2 diabetes (τb=0.365, p<0.001). Those with higher attitude scores were also significantly more likely to agree that this genetic testing should be available through the NHS (τb=0.34, p<0.001).

After Bonferroni adjustment for multiple comparisons (αadj= 0.010), the positive associations between attitude scores and both willingness (p < 0.001) and agreement for NHS provision (p < 0.001) remained statistically significant.

Participants were also asked to what their overall perception of medical research is with 63 (60.6%) indicating it was very positive, 33 (31.7%) indicating it was somewhat positive, 7 (6.7%) indicating they felt neutral and 1 (1.0%) indicating they felt somewhat negative. None of our participants selected “Very negative”. Those who had previous experience of participating in medical research/knew someone who had participated were significantly more likely to have a positive perception of research (τb=0.201, p=0.027).

### Influence on lifestyle

Participants were asked if knowing their risk of developing type 2 diabetes following gestational diabetes would influence their lifestyle (e.g. exercise patterns, diet etc.). 63 (56.3%) answered “Yes, definitely”, 41 (36.6%) answered “Yes, probably”, 5 (4.5%) answered “Not sure” and 3 (2.7%) answered “No, it probably would not”.

Participants’ age was significantly associated with whether knowing their risk of diabetes would change their lifestyle (τb =0 .03, *p* = 0.015), with younger participants being more likely to indicate that this knowledge would lead them to change their lifestyle. This measure was not associated with participants’ education group (p=0.85) or ethnicity grouping, White and Non-White (p=0.109).

Participants were asked to indicate how their behaviour might change if a genetic test during pregnancy showed they had a high chance of developing diabetes in the future (Figure 1). Participants were significantly more motivated to eat a healthy, balanced diet than to take preventative medication (*Z* = 4.35, *p* < 0.001). However, they were significantly more motivated to take medication than to exercise regularly (*Z* = –3.62, *p* < 0.001). There was no significant difference between motivation to exercise regularly and motivation to eat a healthy diet (*Z* = 1.41, *p* = 0.157). This suggests that participants prioritise dietary changes over taking medication and taking medication over exercise but have no preference for dietary changes compared to exercise.

**Figure 1.**
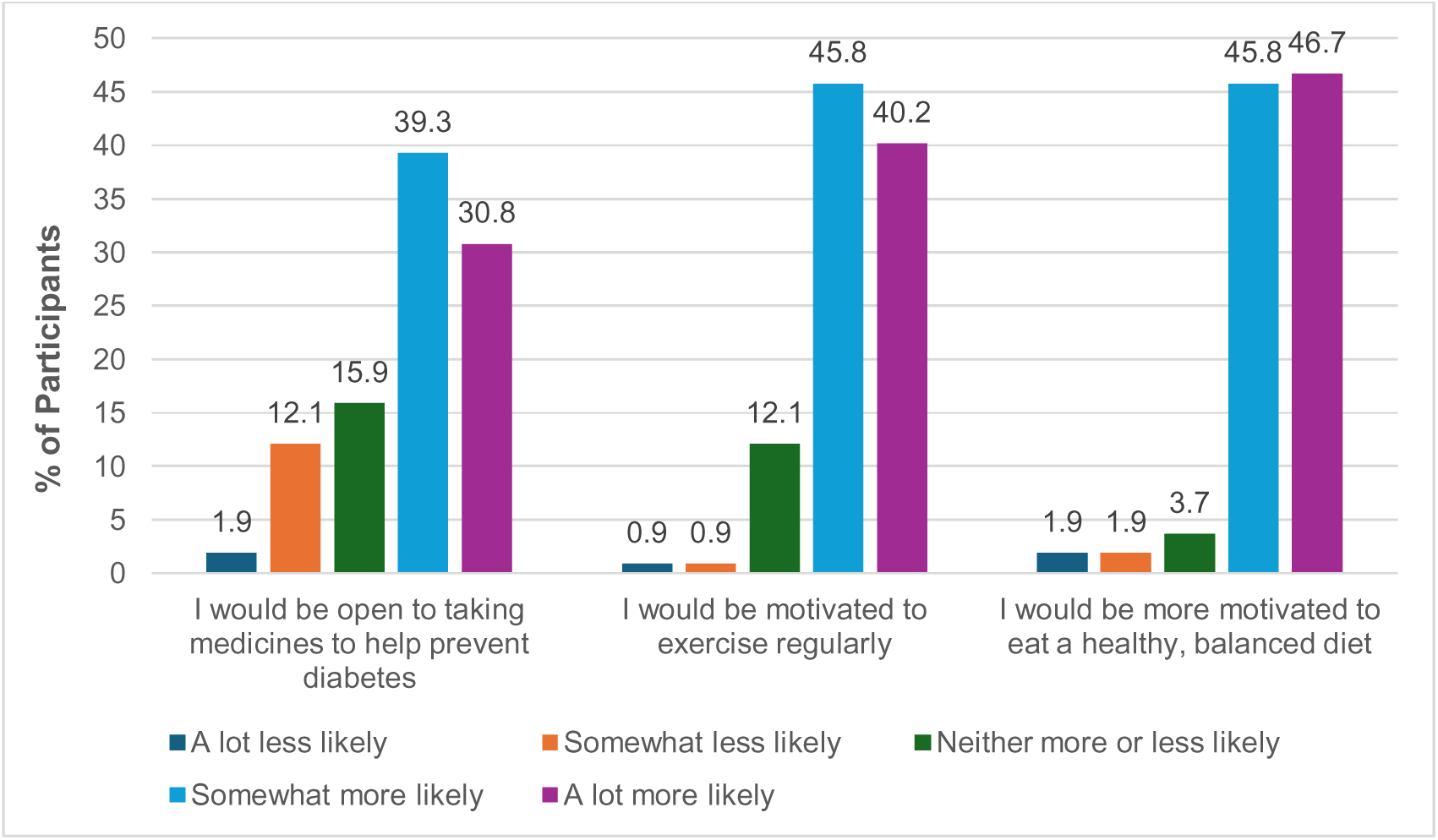
Motivation for lifestyle changes following risk result.

The motivation to both exercise regularly and eat a healthy, balanced diet significantly differed across education groups (F(2,91) = 4.54, p = 0.013, and F(2,91) = 4.42, p = 0.015 respectively). Post hoc Tukey tests indicated that participants with medium levels of education reported significantly higher exercise (M = 4.48) and diet motivation (M = 4.55) than those with low levels of education (M = 3.70 and 3.80, respectively). No significant differences were observed between the medium and high education groups. Motivation to take preventative medications did not significantly differ across education groups (p=0.720).

There were no significant relationships between participants’ age or ethnicity group (White vs. Non-White) and their motivation to exercise (*p* = 0.296 and 0.075, respectively), motivation to make dietary changes (*p* = 0.335 and 0.192), or willingness to take medication (*p* = 0.075 and 0.357).

After Bonferroni correction, the association between participants’ age and likelihood of changing their lifestyle (τb = 0.03, p = 0.015) remained significant (α_adj = 0.0167). For motivation-related comparisons, both the differences between eating a healthy diet and taking preventative medication (Z = 4.35, p < 0.001) and between taking medication and exercising (Z = –3.62, p < 0.001) remained significant after Bonferroni adjustment across the three behaviour comparisons (α_adj = 0.0167). The differences in motivation to exercise and eat a healthy diet across education groups (F(2,91) = 4.54, p = 0.013; F(2,91) = 4.42, p = 0.015) also remained significant within the education-related motivation family (α_adj = 0.0167).

### Concerns about genetic testing

Participants were asked about their level of concern regarding potential uses of data collected from genetic testing (Figure 2). They were most worried about their privacy being compromised and least worried about their ethnicity being identifiable from the data. Overall, participants reported relatively low levels of concern, with a mean score of 9.15 (SD = 3.77, range 6–20), where higher scores indicate greater worry.

**Figure 2.**
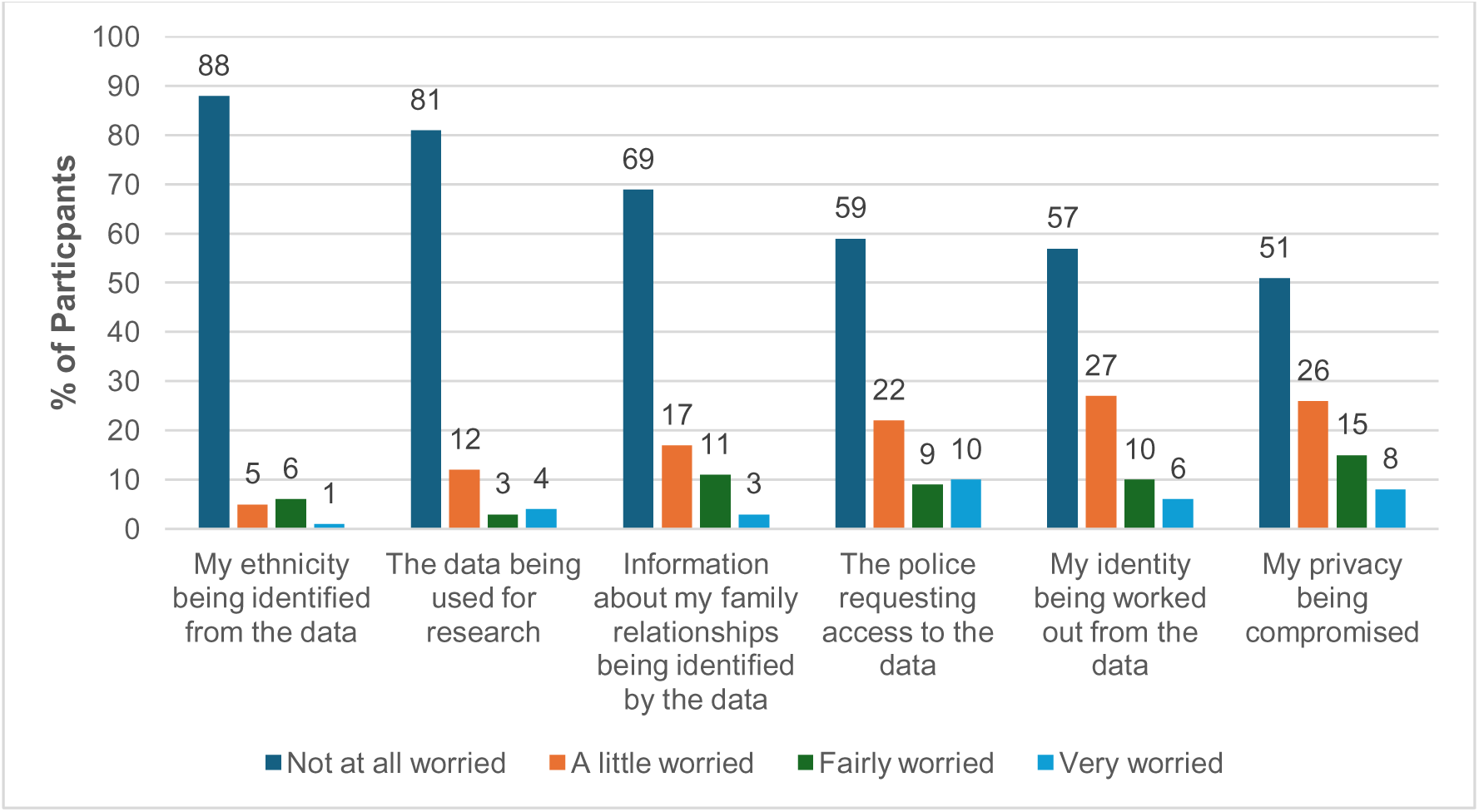
Concerns over use of data following genetic testing.

There was a statistically significant association between participants’ overall concern scores and perceptions of medical research (*H*(3) = 20.05, *p* < 0.001, η² = 0.18). Post hoc pairwise comparisons with Bonferroni correction showed that participants who reported a very positive perception of medical research were significantly less worried about potential uses of the data compared to those with a somewhat positive perception (*p* = 0.001) and those who were neutral (*p* = 0.037). No significant differences were observed between other groups.

Overall, concern scores were not significantly associated with education groups (p=0.915), ethnicity groups, White and Non-White (p=0.531) or age (p= 0.526), suggesting that participants’ low levels of concerns about uses of genetic data were generally consistent across the demographic groups.

Free text responses regarding other concerns around genetic testing were received from 22 respondents. Six themes were identified (Table 2).

**Table 2.**
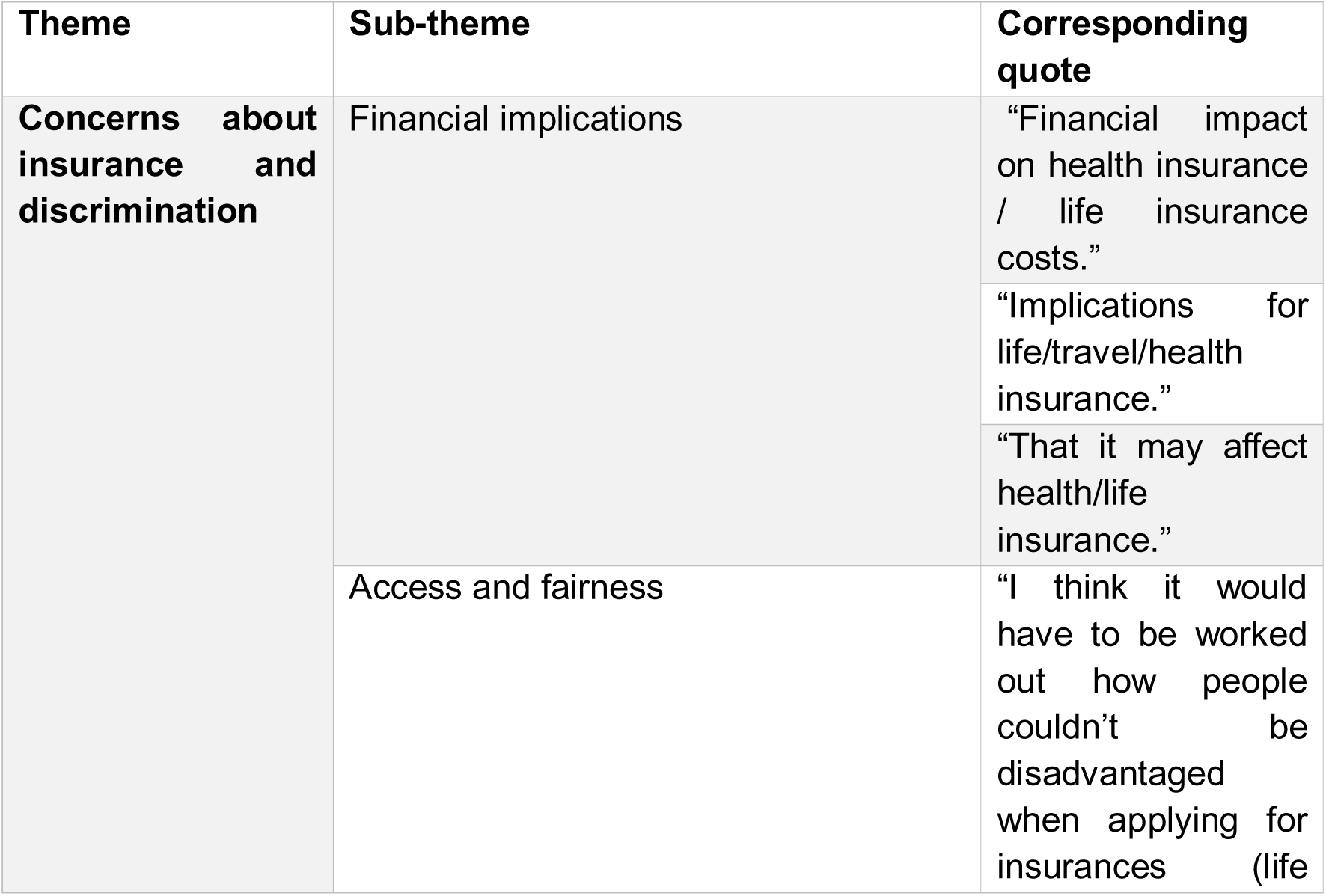
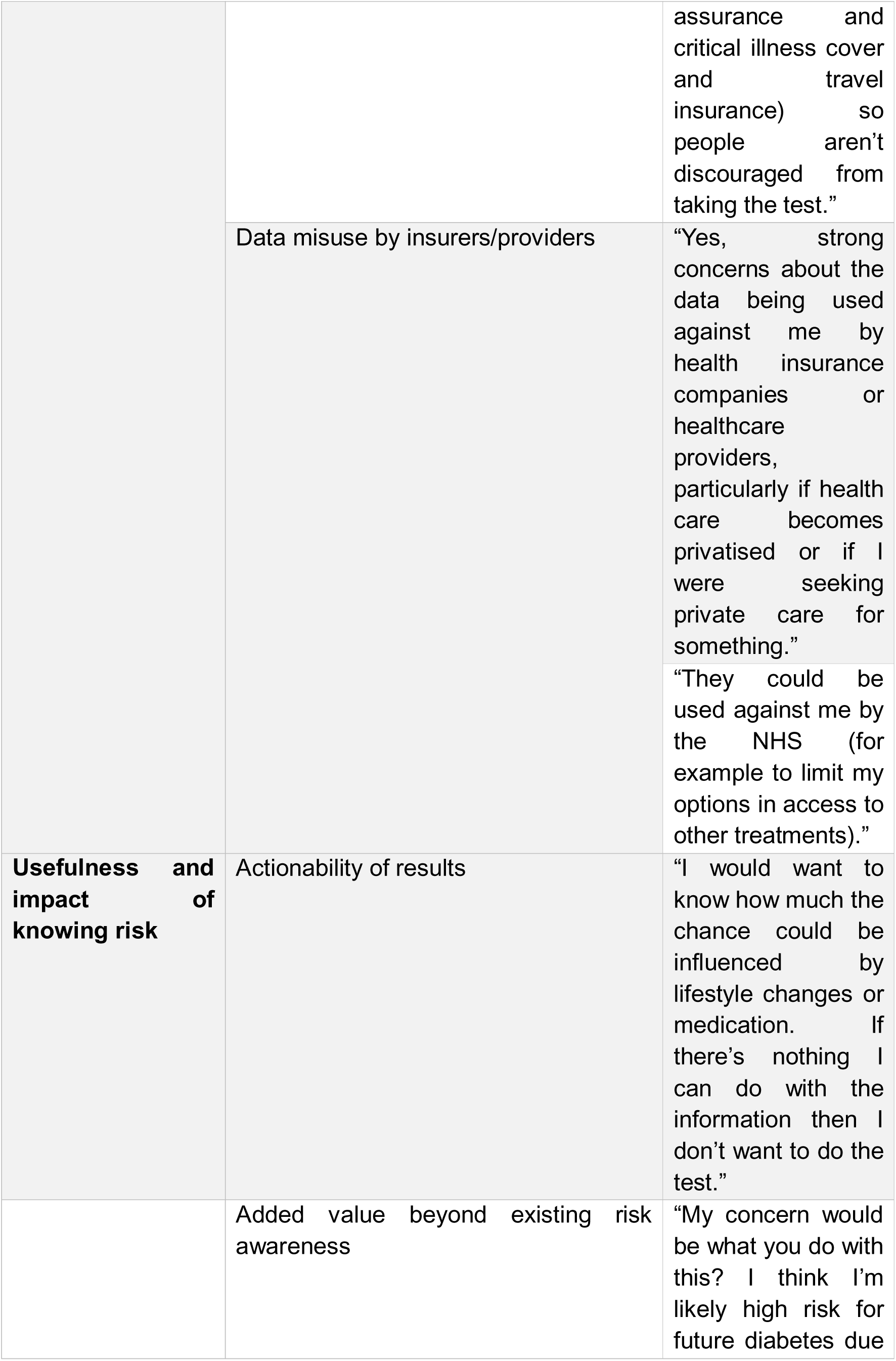
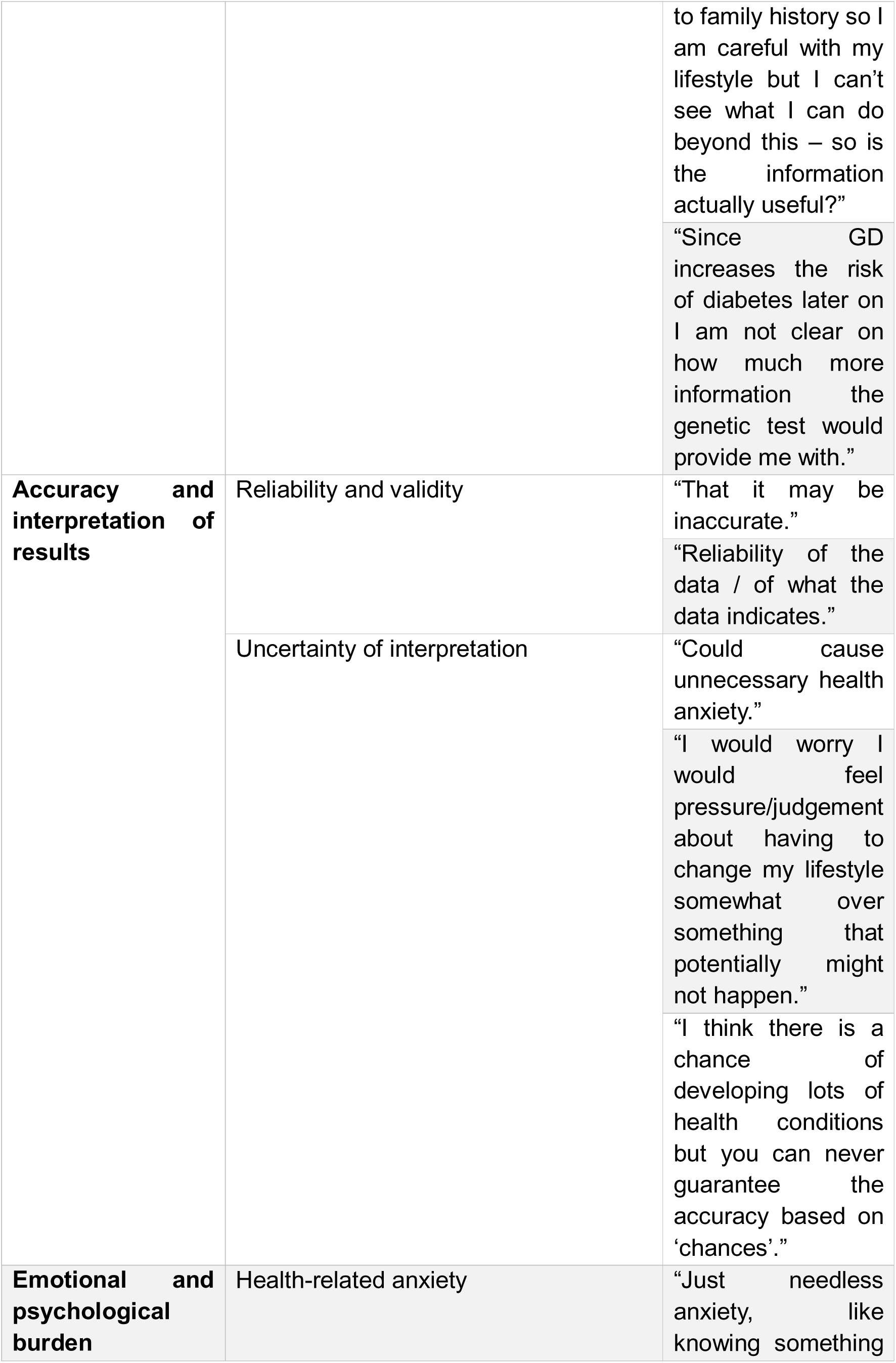
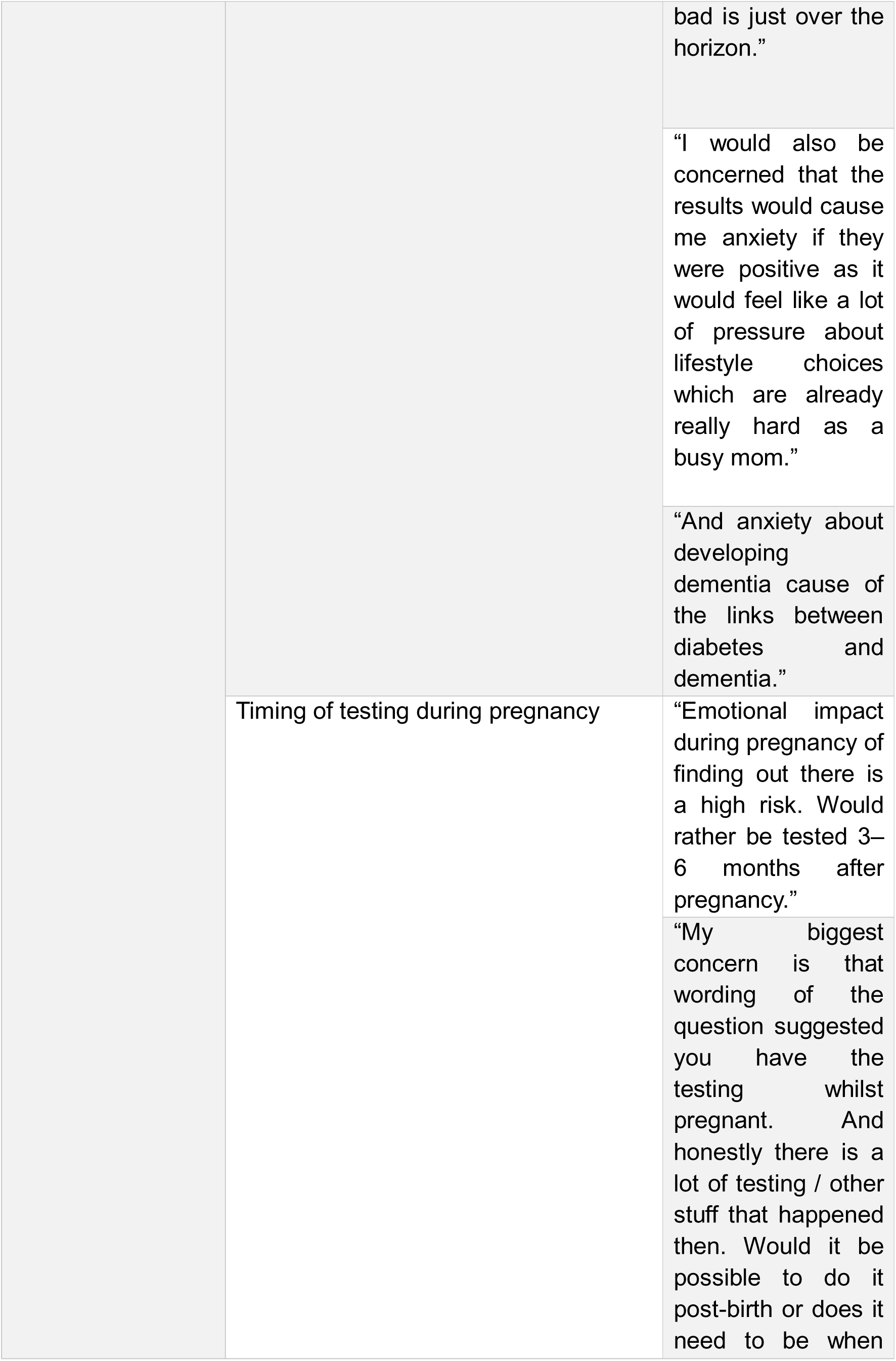
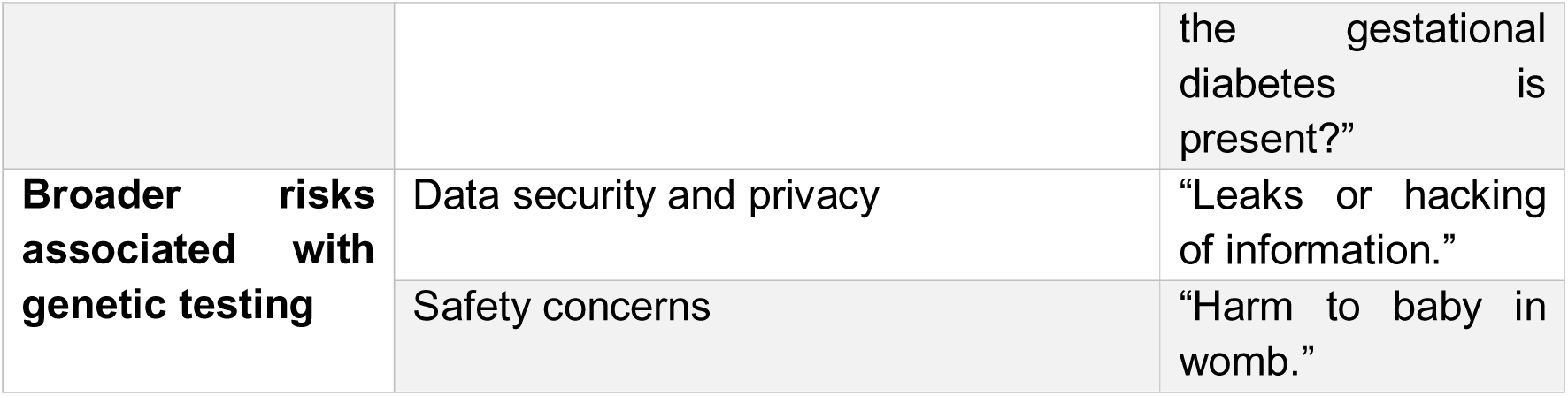
Key themes of participants’ concerns around genetic data collected.

## Discussion

This study explored the acceptability of genetic testing to assess future type 2 diabetes risk among women with current or previous gestational diabetes mellitus, alongside their attitudes towards genetic testing, potential impact of test results on lifestyle, and concerns around data collected.

### Willingness to undergo testing

Overall, our findings indicate high willingness to undergo both non-genetic and genetic testing to assess their risk of future T2DM, with more than 80% of participants reporting willingness to take a genetic test. Importantly, participants were not significantly more likely to favour a simple a non-genetic test over a genetics-based test. Willingness did not differ significantly by participants’ age or education, however, White participants reported greater willingness to take a genetic test and stronger agreement that such testing should be available on the NHS compared with Non-White participants. These findings suggest that while the acceptability of genetic testing was comparable to the acceptability of non-genetic testing for our participants, willingness to engage with genetic testing initiatives differed by ethnicity. This aligns with previous research that has found minority groups to have lower acceptability and greater distrust of genetic testing, potentially due to complex factors such as historical inequities, or concerns about discrimination (17, 18).

Positive attitudes toward genetic testing were also associated with greater willingness to undergo testing and stronger support for its availability through the NHS, indicating that favourable perceptions may directly help engagement with this genetic testing. These findings support evidence that providing clear information and education about genetic testing, its purpose, and potential benefits may increase acceptability and reduce decisional conflict regarding genetic testing (15).

### Influence of results on future lifestyle behaviours

The survey also investigated the potential influence of knowing future T2DM from genetic testing on participants’ lifestyle behaviours. Most participants reported that knowing their future diabetes risk would influence their diet and exercise patterns, with younger participants indicating stronger motivation to change their lifestyle. Motivation to adopt dietary changes was higher than motivation to take preventive medications, and participants with medium levels of education reported higher motivation for diet and exercise than those with low levels of education. These findings suggest that genetic risk information may serve as a catalyst for lifestyle modification and highlight the need for interventions that support behavioural changes following risk disclosure. Participant demographics that reported lower levels of motivation (older participants and those with lower levels of formal education) should be effectively supported through tailored interventions to incorporate sustainable and manageable lifestyle modifications.

### Concerns surrounding testing

Participants who held less positive views of medical research reported comparatively greater worries about data collection and privacy, suggesting that distrust or uncertainty may pose barrier for some. Targeted engagement with these groups could improve understanding of their perspectives, provide an opportunity for public education about genetic risk prediction testing and inform program design to widen engagement.

Although overall worry about data use was low, a small subset of respondents (N=21 out of 112 respondents) provided free-text responses that offered actionable nuance. Recurrent themes included: potential implications for health insurance (echoing broader discourse on genetic discrimination), questions about the actionability and interpretation of results, and the emotional load of testing, especially during pregnancy. These points underscore the importance of clear communication and counselling to ensure that individuals understand what genetic risk information means and how it can guide preventive strategies after GDM.

Taken together, the qualitative analysis, while based on a limited number of comments, highlights specific issues that could influence uptake. Addressing them through transparent data-governance messaging, practical examples of actionability, optional timing (antenatal vs postpartum), and access to psychosocial support may promote equitable and informed participation.

### Implications for future practice

These findings have important implications for clinical practice and policy. Genetic testing for type 2 diabetes risk could be integrated into post-GDM care to support early identification of high-risk individuals and promote preventive lifestyle behaviours. Given the observed demographic differences in acceptability, healthcare providers should consider inclusive approaches, ensuring that information is accessible and that any concerns around privacy, discrimination, and data use are explicitly addressed. Additionally, interventions may benefit from emphasising actionable outcomes from genetic testing, particularly regarding diet and exercise, as these were the areas where participants expressed the strongest motivation for behaviour change.

## Strengths and Limitations

To our knowledge, this is the first study to explore UK women’s perceptions of genetic testing to assess future T2DM risk following GDM. This study combined quantitative and qualitative data, providing both breadth and depth in understanding the perspective of women with experience of GDM toward genetic testing to assess future T2DM risk. The involvement of participants with both current and previous GDM enhances the generalisability of findings within this clinically relevant population.

A key limitation of this work is that our sample was predominantly White and fairly well educated, limiting generalisability to more ethnically and socioeconomically diverse populations. Responses were self-reported and hypothetical, which may introduce social desirability bias and may not reflect actual behaviours if testing were available.

## Future research

Future research should aim to engage larger, more ethnically and socioeconomically diverse samples to better understand potential disparities in acceptability and motivation. Additionally, further qualitative work such as focus groups or interviews, could explore participants’ concerns in greater depth, particularly regarding emotional and psychological impacts, timing of testing, and trust in healthcare systems.

## Conclusion

Women with current or previous GDM demonstrate high willingness to undergo genetic risk-prediction for T2DM, positive attitudes toward genetic research, and generally low concern about data use. Ethnicity was associated with willingness to undergo genetic risk-prediction and support for its NHS availability. Younger participants showed greater motivation to change lifestyle behaviours in response to risk information. These findings support integrating of genetic risk-prediction into post-GDM care, with clear education, behaviour change support, and targeted reassurance on data use to ensure equitable uptake.

## Data Availability

Anonymised data collected through the present study are available upon reasonable request to the authors

## Acknowledgements

The authors would like to thank Jinty Moffett and the interdisciplinary GDM research panel (J Hirst, R Lindsay, R Holt, L Magee, R Reynolds) for their assistance with the survey design. We would also like to thank Diabetes UK, ICP support, the Diabetes Research & Wellness Foundation for sharing this survey and our respondents for their participation.

## References

1. Paulo MS, Abdo NM, Bettencourt-Silva R, Al-Rifai RH. Gestational Diabetes Mellitus in Europe: A Systematic Review and Meta-Analysis of Prevalence Studies. Frontiers in Endocrinology. 2021;12.

2. Bellamy L, Casas JP, Hingorani AD, Williams D. Type 2 diabetes mellitus after gestational diabetes: a systematic review and meta-analysis. Lancet. 2009;373(9677):1773-9.

3. Vounzoulaki E, Khunti K, Abner SC, Tan BK, Davies MJ, Gillies CL. Progression to type 2 diabetes in women with a known history of gestational diabetes: systematic review and meta-analysis. BMJ. 2020;369:m1361.

4. Butalia S, Barrett O, Savu A, Liyanage V, Senior P, Yeung RO, et al. Postpartum Diabetes Screening and Conversion Rates Among Women Diagnosed with Gestational Diabetes Mellitus Using Standard versus Modified Criteria During the COVID-19 Pandemic. Canadian Journal of Diabetes.

5. Kim C, Tabaei BP, Burke R, McEwen LN, Lash RW, Johnson SL, et al. Missed opportunities for type 2 diabetes mellitus screening among women with a history of gestational diabetes mellitus. Am J Public Health. 2006;96(9):1643–8.

6. Zöllner J, Orazumbekova B, Hodgson S, van Heel DA, Akhtar S, Anwar M, et al. Understanding the potential contribution of polygenic risk scores to the prediction of gestational and type 2 diabetes in women from British Pakistani and Bangladeshi groups: a cohort study in Genes and Health. AJOG Global Reports. 2025;5(2):100457.

7. Likhanov M, Zakharov I, Awofala A, Ogundele O, Selita F, Kovas Y, et al. Attitudes towards genetic testing: The role of genetic literacy, motivated cognition, and socio-demographic characteristics. PLoS One. 2023;18(11):e0293187.

8. Aro AR, Hakonen A, Hietala M, Lönnqvist J, Niemelä P, Peltonen L, et al. Acceptance of genetic testing in a general population: age, education and gender differences. Patient Education and Counseling. 1997;32(1):41–9.

9. Niyibizi JB, Rutayisire E, Mochama M, Habtu M, Nzeyimana Z, Seifu D. Awareness, attitudes towards genetic diseases and acceptability of genetic interventions among pregnant women in Burera district, Rwanda. BMC Public Health. 2023;23(1):1961.

10. Kvaratskhelia E, Chokoshvili D, Kvintradze M, Surmava S, Dzagoevi K, Borry P, et al. Public attitudes towards the genetic testing in Georgia. Journal of Community Genetics. 2021;12(3):407–14.

11. Dar-Nimrod I, Heine SJ. Genetic essentialism: on the deceptive determinism of DNA. Psychol Bull. 2011;137(5):800–18.

12. Frieser MJ, Wilson S, Vrieze S. Behavioral impact of return of genetic test results for complex disease: Systematic review and meta-analysis. Health Psychol. 2018;37(12):1134–44.

13. Sendas MV, Freitas MJ. “The needs of women in the postpartum period: A scoping review.”. Midwifery. 2024;136:104098.

14. Sanderson SC, Lewis C, Hill M, Peter M, McEntagart M, Gale D, et al. Decision-making, attitudes, and understanding among patients and relatives invited to undergo genome sequencing in the 100,000 Genomes Project: A multisite survey study. Genetics in Medicine. 2022;24(1):61–74.

15. Patel R, Friedrich B, Sanderson SC, Ellard H, Lewis C. Parental knowledge, attitudes, satisfaction and decisional conflict regarding whole genome sequencing in the Genomic Medicine Service: a multisite survey study in England. Journal of Medical Genetics. 2025;62(4):289–97.

16. Thomas DR. A General Inductive Approach for Analyzing Qualitative Evaluation Data. American Journal of Evaluation. 2006;27(2):237–46.

17. Harris BHL, McCabe C, Shafique H, Lammy S, Tookman L, Flanagan J, et al. Diversity of thought: public perceptions of genetic testing across ethnic groups in the UK. Journal of Human Genetics. 2024;69(1):19–25.

18. Suther S, Kiros G-E. Barriers to the use of genetic testing: A study of racial and ethnic disparities. Genetics in Medicine. 2009;11(9):655–62.

19. Lou S, Lomborg K, Lewis C, Riedijk S, Petersen OB, Vogel I. “It’s probably nothing, but…” Couples’ experiences of pregnancy following an uncertain prenatal genetic result. Acta Obstetricia et Gynecologica Scandinavica. 2020;99(6):791–801.

