## Appendix A for "Understanding Patient Perceptions of Genetic Testing to Predict Type 2 Diabetes Risk After Gestational Diabetes"

Appendix A- Patient Survey

Start of Block: Introduction

Introduction

**You are invited to share your thoughts on genetic testing and chance of developing future diabetes.** We want to hear your views on genetic testing.
This research focuses on understanding the future chance of type 2 diabetes after gestational diabetes. During this survey when we ask about 'diabetes' we are referring to type 2 diabetes.

**Why is this research important?**
Some women develop gestational diabetes during pregnancy (having too much sugar in your blood). It affects about 6% (around 40,477) of pregnancies in the UK each year.
Gestational diabetes can lead to health complications during pregnancy and birth and increases the risk of developing type 2 diabetes later.

We are exploring how genetic testing could help to predict who is more likely to develop type 2 diabetes after having gestational diabetes. We want to understand your thoughts on this genetic testing, whether you would want to know your future risk of developing type 2 diabetes and how this might influence your health choices. Your feedback will help researchers and healthcare providers better understand attitudes toward genetic testing.

**Why have I been invited to take part?**
You have personal experience with gestational diabetes, either now or in the past.
While you may not personally benefit from the results, your answers could help improve care for other women in the future.

**What will I need to do?**
If you would like to take part, simply tick the consent below and complete the short questionnaire (attached). There are no extra tests or visits involved.
The questionnaire will take about 5 minutes to complete. Do not worry if you cannot answer all of the questions.

**Do I have to take part?**
No, you do not have to take part if you do not want to. Participation is entirely voluntary, and it is up to you to decide if you would like to do so.

**How will my information be used?**
All the responses will be kept strictly confidential and anonymous. No one will be able to identify you from the results. The data will be stored on a UCL database and only accessible to the research team.

**Want to know more?** If you have any questions, feel free to contact us at.
**Ready to take part?** Please tick the consent box below and complete the questionnaire. If

you are happy to be contacted about future research relating to this questionnaire, you can also leave your phone number or email address.

☐ I acknowledge that I have read and understood the above information. I consent to participate in this study. (1)

☐ I consent to be contacted for follow up focus groups. (If you agree, please enter your email/phone number below) (2)

________________________________________________

Q1 Please tick which applies

○ I currently have gestational diabetes (1)

○ I have previously had gestational diabetes (2)

○ I have never been diagnosed with gestational diabetes (3)

**End of Block: Introduction**

**Start of Block: Attitudes towards genomic testing**

DNA is a unique code you inherit from your parents that carries the information needed for your body to grow and function. It remains the same throughout your life. Your DNA can influence your chance of developing diseases like diabetes.

By understanding variations in your DNA - also known as genetics – you may learn whether you have a higher or lower chance of developing diabetes. A simple genetic test using saliva or blood can provide doctors with a clearer idea of your individual chance. This information helps them to offer a tailored intervention programme, a process known as personalised medicine.

Q2 If you were recommended to have a test (not based on genetics) to find out your chance of developing diabetes in the future would you be willing to take it?

- ○ Yes (1)
- ○ No (2)
- ○ Not sure (3)

---

Q3 If you were recommended to have a simple genetic test (a blood test) to find out your chance of developing diabetes in the future would you be willing to take it?

- ○ Yes (1)
- ○ No (2)
- ○ Not sure (3)

---

Q4 On a scale of 1 to 5, please rate how you feel about having a genetic test during pregnancy to find out your chance of developing diabetes later in life:

| | 1 | 2 | 3 (neutral) | 4 | 5 | |
|---|---|---|---|---|---|---|
| | 1 (1) | 2 (2) | 3 (3) | 4 (4) | 5 (5) | |
| Harmful | ○ | ○ | ○ | ○ | ○ | Beneficial |
| Unimportant | ○ | ○ | ○ | ○ | ○ | Important |
| A bad thing | ○ | ○ | ○ | ○ | ○ | A good thing |
| Not helpful | ○ | ○ | ○ | ○ | ○ | Helpful |

---

Q5 If a genetic test during pregnancy indicated you had a high chance of developing diabetes in the future, would this information influence your lifestyle (e.g. exercise patterns, diet etc.)?

- ○ Yes, definitely (1)
- ○ Yes, probably (2)
- ○ Not sure (3)
- ○ No, it probably would not (4)
- ○ No, it definitely would not (5)

---

Q6 Please use the scales below to indicate how your behaviour might change if a genetic test during pregnancy showed you had a high chance of developing diabetes in the future.

| | A lot less likely (1) | Somewhat less likely (2) | Neither more nor less likely (3) | Somewhat more likely (4) | A lot more likely (5) |
|---|---|---|---|---|---|
| I would be more motivated to exercise regularly (1) | ○ | ○ | ○ | ○ | ○ |
| I would be more motivated to eat a healthy, balanced diet (2) | ○ | ○ | ○ | ○ | ○ |
| I would be open to taking medicines to help prevent diabetes (3) | ○ | ○ | ○ | ○ | ○ |

---

Q7 Imagine the NHS offered genetic testing to predict your chance of developing diabetes. How much do you agree or disagree that this testing should be available?

- ◯ Strongly agree (1)
- ◯ Agree (2)
- ◯ Neither agree nor disagree (3)
- ◯ Disagree (4)
- ◯ Strongly disagree (5)

**End of Block: Attitudes towards genomic testing**

---

**Start of Block: Concerns About Genetic Testing for Diabetes Risk**

Q8 Thinking about genetic testing to predict your future chance of developing diabetes after gestational diabetes, how worried, if at all, would you be about the following things? Please select one answer for each statement.

| | Not at all worried (1) | A little worried (2) | Fairly worried (3) | Very worried (4) |
|---|---|---|---|---|
| My privacy being compromised (1) | ○ | ○ | ○ | ○ |
| My identity being worked out from the data (2) | ○ | ○ | ○ | ○ |
| My ethnicity being identified from the data (3) | ○ | ○ | ○ | ○ |
| Information about my family relationships being identified from the data (4) | ○ | ○ | ○ | ○ |
| The data being used for research (5) | ○ | ○ | ○ | ○ |
| Police requesting access to the data (6) | ○ | ○ | ○ | ○ |

---

Q9 Do you have any other concerns about taking a genetic test to predict your chance of developing diabetes in the future, the results, or how they might be used? (Please specify)

________________________________________________________________

Q10 If genetic testing data were used to improve diabetes care and research, which, if any, of the following groups would you be comfortable sharing your genetic data with for research purposes? Please select all that apply.

- ☐ Healthcare professionals (e.g., doctors, nurses, and pharmacists) (1)
- ☐ The national medicine regulator (the agency responsible for ensuring medicines are safe and effective) (2)
- ☐ Academics or university-associated researchers (3)
- ☐ Charities or other non-profit organisations (4)
- ☐ Private companies (pharmaceutical or biotech firms) (5)
- ☐ None of the above (6)

---

Q11 Have you ever had any pre-natal genetic testing? e.g Chorionic Villus Sampling, Amniocentesis, NIPT or exome sequencing

- ○ Yes- If you are happy to specify please do so below. (1) ________________________________________________
- ○ No (2)

End of Block: Concerns About Genetic Testing for Diabetes Risk

---

Start of Block: Attitudes towards research

Q12 What is your overall perception of medical research?

- Very positive (1)
- Somewhat Positive (2)
- Neutral (3)
- Somewhat Negative (4)
- Very negative (5)

---

Q13 Have you ever taken part in research, or do you know someone who has?

- Yes, both (1)
- Yes, I have taken part only (2)
- Yes, I know someone who has (3)
- No (4)
- Prefer not to say (5)

**End of Block: Attitudes towards research**

---

**Start of Block: Participant characteristics**

Q14 What is your age?

________________________________________________________________

---

Q15 Which of these best describes your ethnic group?

- ○ Asian, or Asian British (1)
- ○ Black, Black British, Caribbean, or African (2)
- ○ Mixed or multiple ethnic groups (3)
- ○ White: British or Irish (4)
- ○ White: Any other White background (5)
- ○ Other: Any other ethnic group (6) ________________________________________________
- ○ Prefer not to say (7)

---

Q16 Which of these is your highest educational qualification?

- ○ No qualification (1)
- ○ GCSE or O Level (High School exams taken at 16 years old) (2)
- ○ GCE, A-level or equivalent (Education and exams taken at 17-18 years old) (3)
- ○ Vocational (BTEC/NVQ/Diploma) (Education and exams taken at 17-18 years old) (4)
- ○ Bachelors degree or equivalent (University degree) (5)
- ○ Masters degree or equivalent (University degree) (6)
- ○ PhD, MD, or equivalent (University degree) (7)
- ○ Prefer not to say (8)
- ○ Other (9) ________________________________________________

---

Q17 What is your postcode?

________________________________________________________________

________________________________________________________________

________________________________________________________________

________________________________________________________________

________________________________________________________________

**End of Block: Participant characteristics**

---

**Start of Block: Final section**

End of the survey Thank you for taking part! If you would like to be contacted about future research related to this survey, please provide your contact details (a telephone number or email) below. Participation in any future research is entirely voluntarily, and there is no obligation to participate.

○ Add your email / telephone here __________________________________________________

---

Q18 If you have any questions, please feel free to contact.

If you would like to find out more about genetic testing and DNA, we recommend the following resource from Genomics England-https://www.genomicsengland.co.uk/genomic-medicine/understanding-genomics

If you would like further support after completing this survey, Diabetes UK offers a dedicated helpline number at 0345 123 2399. This service is available for people with diabetes, their family or friends, or anyone concerned about their chance of developing diabetes.

**End of Block: Final section**

---
